# Widespread annual rhythms in pediatric emergencies

**DOI:** 10.1101/2024.12.18.24319175

**Authors:** Patricia Tachinardi, Rochelle M Witt, Gang Wu, Jiffin K Paulose, Bala S C Koritala, Hector R Wong, Eneida A Mendonca, David F Smith, John B Hogenesch, Marc D Ruben

## Abstract

Assessments of emergency department (ED) utilization for specific medical conditions reveal distinct annual rhythms, providing valuable insights into risk factors and optimal clinical staffing. However, focusing on a single condition in isolation can lack essential context. Such rhythms may (i) depend on co-occurrence with other conditions, (ii) be obscured by systemic factors that influence all conditions similarly, or (iii) offer limited clinical utility without understanding their rhythmic effect sizes relative to other emergencies. Using a unified framework for analysis, we studied the annual variation in incidences of all chief complaints (CCs) from 1.5 million admissions to our pediatric ED and urgent care centers from 2010 to 2021, enabling comparison of seasonality, effect sizes, and interactions across all types of emergencies. Most CCs (∼80%) had annual rhythms, with a range of phases. Specific patterns, such as spring and fall peaks in neurologic-, injury-, and psychiatric-related CCs, have immediate significance. For example, psychiatric emergencies, which the American Academy of Pediatrics has designated a national crisis, were among the largest rhythmic effect sizes of all CCs. Further evaluation integrating ICD-10 diagnoses identified patient subtypes for psychiatric and other CCs, suggesting distinct annual influences. Using counterpart data from across Brazil, we identified marked global differences in annual patterns of ED utilization, including psychiatric emergencies. Lastly, we identified CCs with large *weekday* effects, impacting care and staffing needs, especially when combined with annual rhythms.

## Introduction

Patients seek care at the emergency department (ED) for diverse medical issues, prompting interest in presentation patterns. Knowledge of predictable changes, or rhythms, in ED utilization can inform staffing, prevention, and insights into causes. Studies of total ED admissions reveal rhythms by time of day (afternoon peak) (1), week (weekend–Monday peak) (2, 3), and year (winter and summer peaks) (2). This knowledge helps to forecast overall ED volume, but falls short in uncovering the impact of individual medical conditions.

Alternatively, studies of ED presentation patterns focus on single medical conditions. For example, descriptions of annual rhythms in asthma (4–6), mental health (7, 8), and other emergencies shed light on risk factors for these conditions. However, studies that focus on a single condition may overlook crucial contextual factors, complicating interpretation. First, rhythms in one condition may depend on the co-occurrence of another condition. For instance, suicidal behavior, a common reason for ED visits, is positively associated with other psychiatric (9–11) and nonpsychiatric conditions (12–15). It is not known how comorbid profiles distinguish ED rhythm profiles. A second limitation of single-condition studies is that broader systemic influences may obscure rhythms specific to a particular condition. For example, medical emergencies seem to correlate with daylight and typical wake hours (1, 16, 17), potentially masking underlying rhythms, such as nocturnal asthma (18). In sum, interpreting rhythms in one condition can be improved by contextualizing with all other conditions.

Computational tools applied to electronic medical records (EMR) data allow analysis of all medical emergencies in a unified framework. We studied the annual variation in incidences of all chief complaints (CCs) from 1.5 million admissions to our pediatric ED and urgent care centers from 2010 to 2021, enabling comparison of annual phases (ie. time of the year when admissions for a given condition peaked), effect sizes, and interactions across emergencies. We focused on the annual time-scale because, despite seasonal influences on human physiology (19–21), time of year is not typically a consideration in healthcare, outside of allergy and infectious disease.

There is also a lack of understanding of the many factors that shape annual patterns in ED presentation. Even in asthma, where seasonality is well studied, exacerbation is loosely understood as a combination of viral, allergenic, social, and intrinsic factors (5, 18). A unified framework may help to infer causality.

We found that most CCs (∼80% of 318 CCs analyzed) in a pediatric ED in Cincinnati, USA, varied by month of the year, with a range of phases. Phases were marked by therapeutic areas. Some of these are well known, for example, peak respiratory-related CCs in the winter, and skin-related CCs in summer. Others were unexpected; for example, peaks in child neurologic-, injury-, and psychiatric-related CCs in the spring and fall, which have immediate significance. Psychiatric emergencies, which the American Academy of Pediatrics recently designated a national crisis (22), were among the most seasonal of all CCs. We next evaluated for hidden subsets of patients within each CC based on additional diagnostic information. For instance, patients with the CC of Psychiatric Evaluation separated into fall-peaking, spring-peaking, and arrhythmic classes based on ICD-10 profiles, suggesting descriptions of psychiatric subtypes with distinct seasonal influences. Lastly, it is known that ED volume increases on certain days of the week. We identified CCs with extreme weekday effects, with implications for hospital preparedness, especially when considered on top of annual rhythms.

Separately, we evaluated annual rhythms in emergency admissions across four Brazilian municipalities, covering various latitudes and climates. Asthma, known for seasonality in Brazil even in locations close to the equator (23, 24), was used to validate the public health database (DataSUS). We then focused on mental health conditions to compare with Cincinnati data.

Annual asthma rhythms were found at all locations, with varying patterns possibly linked to environmental factors. For mental health, significant annual rhythms, with differences from Cincinnati, were detected in three locations, suggesting important social and cultural influences. Code to reproduce all analyses is available at https://zenodo.org/records/14502802.

## Results

### Reasons for emergency visits vary by time of year

Our Cincinnati dataset included all 1,545,630 admissions to our institution’s emergency department and five affiliated urgent care centers from January 2010 to December 2021 (fig. S1A, see “*Study cohort*” section in Methods). A patient’s chief complaint (CC) describes the main reason why they seek medical attention. We modeled smooth annual variation in the incidences of CCs with at least 500 admissions (see “*Annual patterns of CCs*” in Methods). 244 of 318 CCs (∼80%) varied by month (FDR < 0.01, chi-square test that smooth term is zero, fig. S1B, File S1) with a range of annual patterns (Fig. 1).

**Fig 1.**
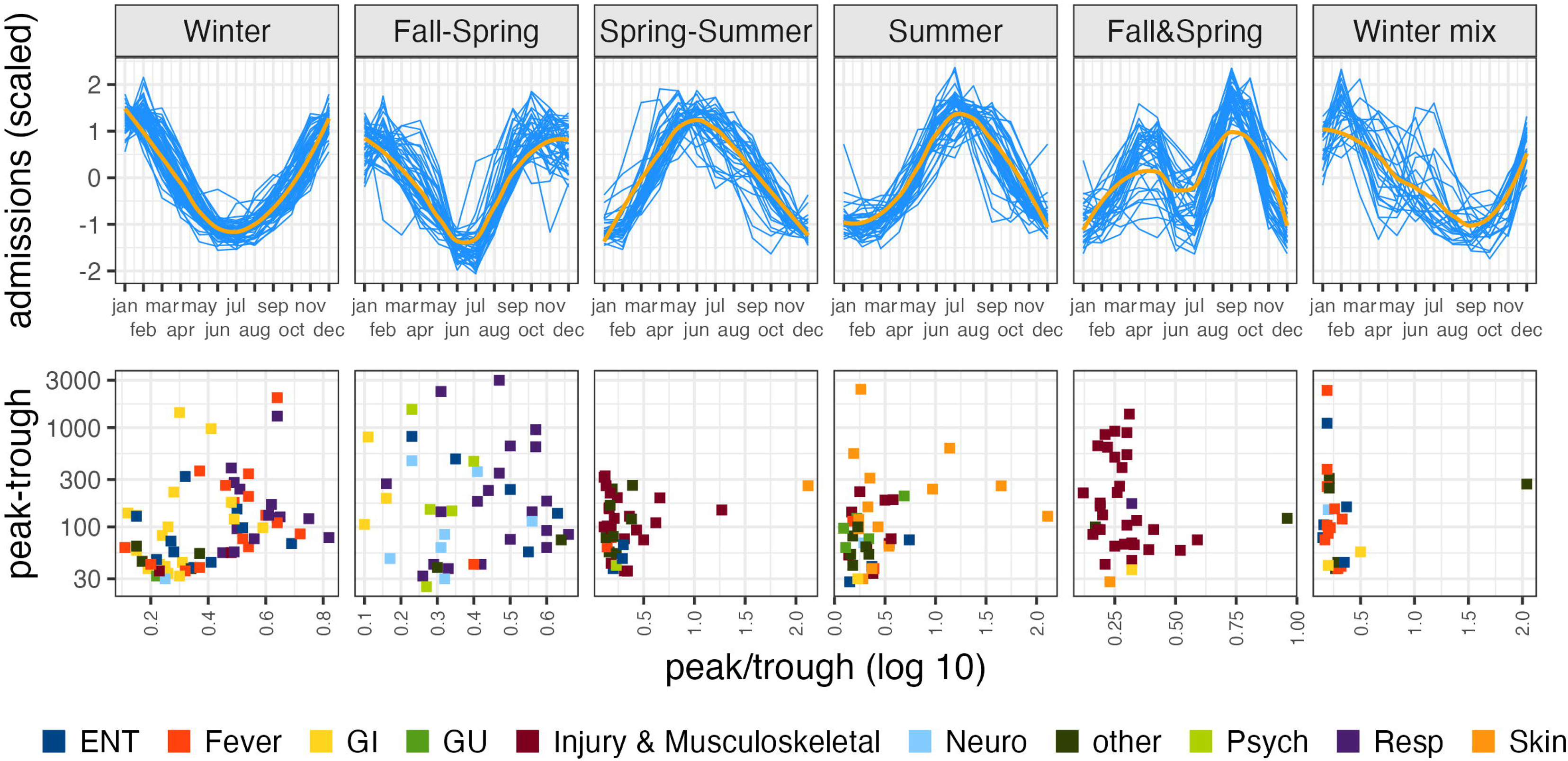
Chief complaints present with annual rhythms. *Top*: K-means clustering partitions the 244 CCs (∼80%) with annual variation (FDR < 0.01, chi-square test that smooth term is zero, fig. S1B) into six groups based on similarities in phase (see “*Rhythm clusters*” section in Methods). Centroids (orange) approximate the overall phases of each cluster. *Bottom:* Squares indicate the effect size and therapeutic area of CCs from the clusters above (see “*Rhythm effect sizes”* section in Methods). Note: x-axis scale varies between clusters to help visualize the spread of peak:trough ratios. *ENT:* ear, nose, throat*; GI:* gastrointestinal*; GU:* genitourinary*; Neuro:* neurological*; Psych:* psychiatric*; Resp:* respiratory.

### Annual phases are marked by therapeutic areas

Studies of annual patterns of illness have focused on single areas, for example asthma (18). Considering all types of medical emergencies in one framework allowed us to compare annual phases, effect sizes, and covariate interactions. Time-of-year clusters are marked by therapeutic areas (Fig. 1, File S1). Some of these are well known, for example peak respiratory-related CCs in the winter, and skin-related CCs in summer. Others were unexpected, for example, peaks in child psychiatric-, neurologic-, and injury-related CCs in both the spring and fall.

Cincinnati, Ohio (39°6’ N and 84°30’W) is positioned away from the equator and sees the full spectrum of seasonal changes in temperature, precipitation, vegetation, and daylight. How these or other variables contribute to annual phases of CCs is unknown. Even in asthma, where seasonality is well studied, exacerbation is loosely understood as a consequence of viral, allergenic, social, and intrinsic factors (18). The range of rhythmic CCs in our dataset, from *Suicidal* to *Migraine* to *Ankle injury* (fig S1C, File S1), suggests a multitude of causes.

### Rhythm effect sizes highlight healthcare burdens

Healthcare impact is dictated by effect sizes, which we consider in two ways (Fig. 1, see “*Rhythm effect size”* in Methods). First, peak-to-trough ratio (PTR) is the fold difference in CC admissions between peak and trough months. PTR is a common measure of change, but it ignores patient volume: 10-to-1 and 10,000-to-1,000 have the same PTR but very different implications. Therefore, to capture patient volume, we consider peak-minus-trough. By this measure, psychiatric emergencies had the largest seasonal effects of all CCs (File S1).

The relationships between month of the year and admission (CC curves in Fig. 1) may depend on factors such as the year, admit age, or biological sex. We tested if our annual model fits were improved by the addition of interaction terms (fig. S2A, see “Covariate interactions” in Methods). Not surprisingly, annual rhythms in the CC *Covid-19* strongly depended on the year because there were no Covid-19 CCs at our institution before 2020. The CC *Coughing and Fever* also depended on the year, due mostly to the deviation in 2021 from expected annual rhythms (fig. S2B). Sex had a small but consistent influence on the annual rhythms of several injury-related CCs (fig. S2C), perhaps due to the nature of fall-time sports (25). Overall, the relationship between month and admissions were minimally affected by sex or year, indicating a robustness of CC annual rhythms throughout the 12 years studied (Fig. 1).

### Hidden patient types and rhythms

Next, we analyzed whether annual rhythms differed across specific patient groups and diagnoses within each CC. Chief complaints differ in complexity (Fig 2A). For some CCs, such as *Foreign Body in Nose*, patients are typically discharged with the same one or two diagnostic (ICD-10) codes. For other CCs, such as *Psychiatric Evaluation*, patients are discharged with multiple ICD-10 codes that can be quite different from one another (Fig. 2A, see ‘CC *patient complexity*’ in Methods). We used this information to better understand patient and rhythm subtypes.

**Fig. 2.**
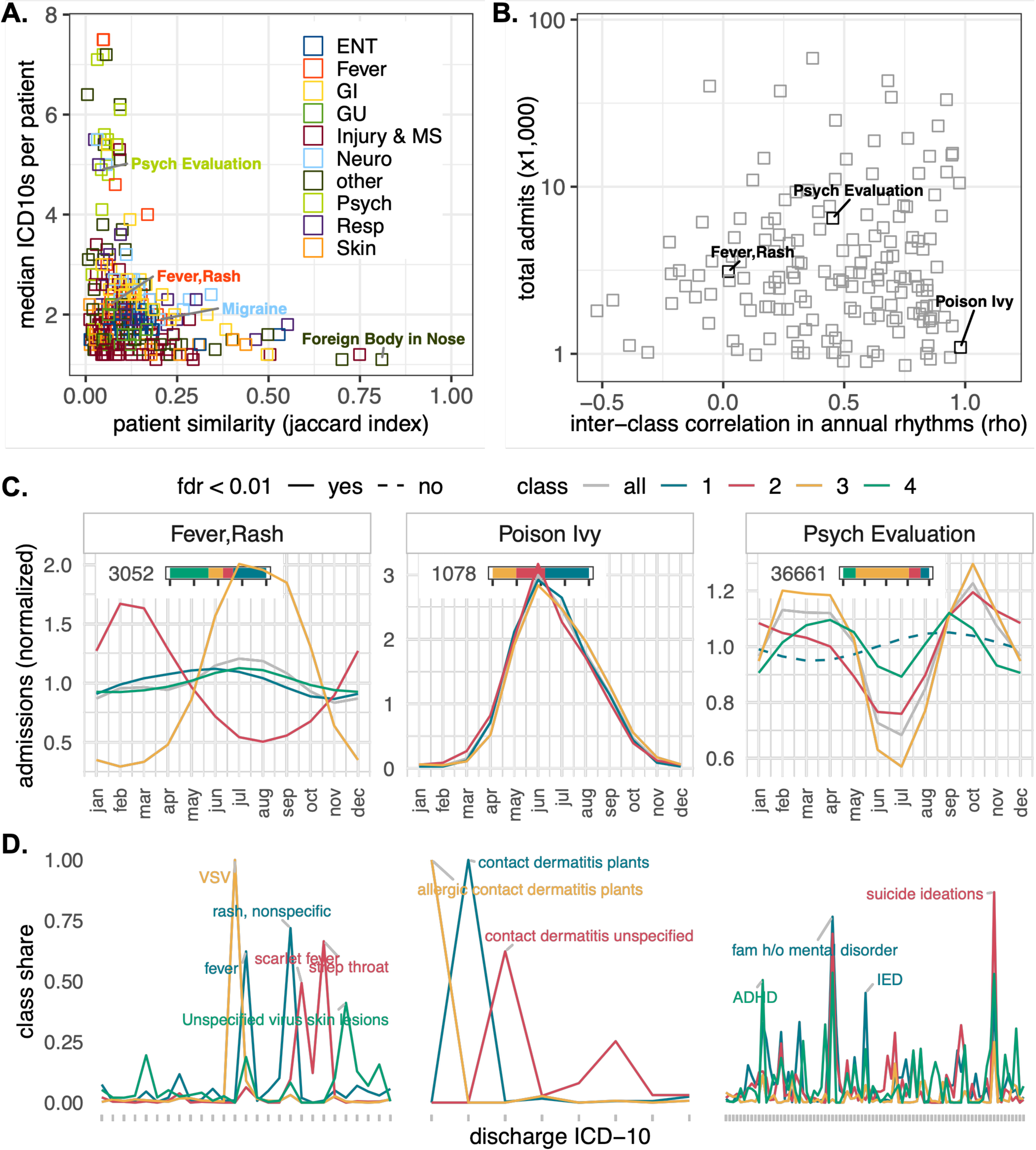
Clinical features identify patient and rhythm subtypes. (**A**) Median number of discharge ICD-10 codes per admission (x-axis) versus the average Jaccard similarity in ICD-10 code makeup between all pairs of admissions (y-axis). A similarity score equal to one indicates that all admissions with that CC had identical discharge ICD-10 code profiles (see “CC *patient complexity*” in Methods). **(B)** LCA identified classes of each CC based on ICD-10 profiles (see “*Latent class analysis”* in Methods). For each CC, we computed the correlation in annual patterns of admission for all possible pairs of classes (see “*Spearman rank correlation”* in Methods). The mean correlation value for each CC (x-axis) represents a single measure of inter-class similarity in annual patterns of admission. A value of 1 indicates a perfect positive correlation, meaning that all pairs of classes for a given CC rose and fell with the same pattern. A value of -1 indicates a perfect negative correlation. **(C)** Annual patterns of admission for each class of three example CCs. Curves were fit as described previously (see “*Annual patterns of CCs”* in Methods). **(D)** The share of total admissions (y-axis) in each class with that ICD-10 (x-axis). The labels and their color indicate the class that is most defined by that ICD-10 code. Labels were limited to ICD-10s with at least 40% share in at least one class. *VSV: Vesicular Stomatitis Virus; ADHD: Attention Deficit Hyperactivity Disorder; IED: Intermittent Explosive Disorder*.

Latent class analysis (LCA) is a method to identify hidden subtypes within a population based on observed categorical variables (30). We applied LCA to identify subtypes, or classes, within each CC based on ICD-10 profiles (see “*Latent class analysis”* in Methods). LCA identified up to four classes for each CC. We then modeled the annual variation in each class, and asked whether different classes had different annual patterns (Fig. 2B, see “*Spearman rank correlation”* section in Methods). For some CCs, for example, *Fever–Rash*, classes had different rhythmic patterns corresponding to different pathogenic sources of infection (Fig. 2C & D). For other CCs, for example *Poison Ivy,* classes had nearly identical rhythms. Most CCs, however, fall somewhere between *Poison Ivy* and *Fever-Rash* (Fig. 2B). For example, *Psychiatric Evaluation* had classes with fall peaks, spring peaks, and no peaks, potentially describing types of psychiatric emergencies with distinct seasonal influences (Fig. 2C & D). Files S2, S3, and S4 detail the ICD-10 makeups and annual profiles for all CC classes. This information has potential implications for hospital staffing, public health intervention, and understanding causes.

### Acute impacts on emergency admissions

So far, we’ve focused on smooth annual changes in admission that we presume are driven by smooth annual changes in daylight, viral prevalence, social schedules, and potentially many other factors. However, there are also *acute* annual changes, for example holidays, first day of school, clock time changes, and other factors that may impact emergency admissions.

To assess acute changes, we calculated 3-day rolling rates of admission changes from Jan 1, 2010, to Dec 31, 2021 (fig. S3, see ’Rolling change’ in Methods). ED and urgent care (UC) admissions were analyzed separately. There is an unmistakable weekly rhythm in admissions, with a Monday peak in the ED, and a Sunday peak in UC (Fig. 3A & B). Monday peaks in the ED have been described (31) but are not universal (2) likely influenced by community factors such as UC weekend availability. Importantly, there were certain CC areas with substantially larger weekly effects (Fig. 3C), which have implications for care and staffing, especially when considered on top of annual rhythms. For instance, psychiatric-related admissions were 67% higher in October compared to June, but 220% higher for Mondays in October compared to Saturdays in June (Fig. 3D).

**Fig 3.**
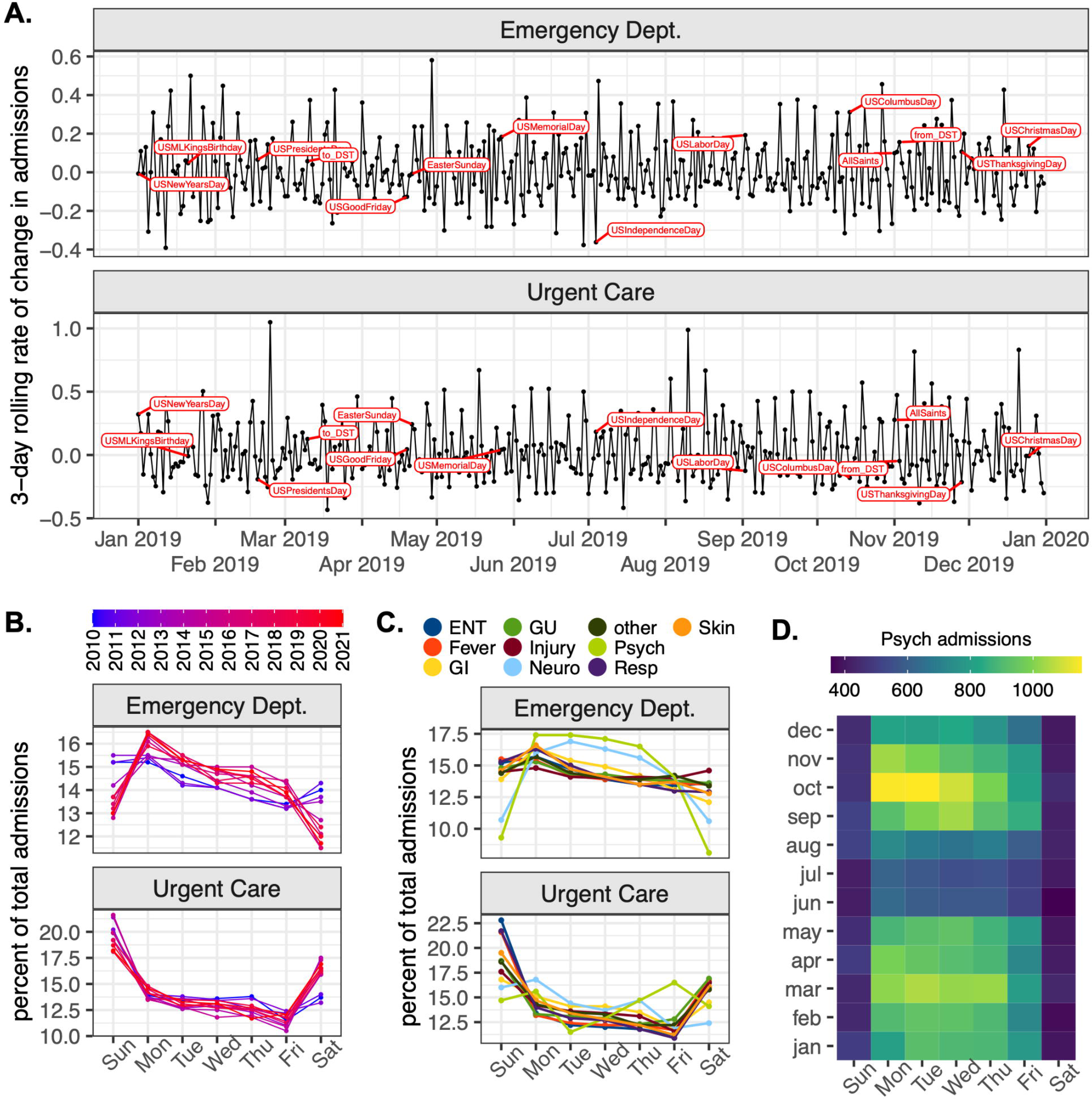
Weekly rhythms in admissions. **(A)** 3-day rolling rates of change in admissions from Jan 01, 2019 to Dec 31, 2020 (see “Rolling change” in Methods). Labels indicate major US holidays and clock time shifts to and from daylight savings time. **(B)** Percent of admissions by day of the week for each year, and **(C)** CC area. *ENT: ear, nose, throat; GI: gastrointestinal; GU: genitourinary; Neuro: neurological; Psych: psychiatric; Resp: respiratory.* **(D)** Total psychiatric-related admissions by month and day for the full study period January 2010 to December 2021.

A second clear pattern emerges from 2010 through 2013. UC admissions appear to spike on the day after major US holidays (fig. S3). This is an artifact of our analysis because prior to 2014 these UC clinics were closed on holidays, yielding spikes in the rolling rate of change for the day after. After 2014, there were no obvious holiday effects (Fig 3A).

Clock time shifts pose another potentially acute effect. Prior studies suggest that injuries and illnesses increase in the days after the springtime shift to daylight savings time (DST) (32). These studies have focused on single conditions, mostly adult, and have not always considered confounding weekly and seasonal effects. We evaluated the specific impact of time shifts, both to and from DST, across all pediatric emergencies. For each CC area, we (1) computed the percent difference in admissions the seven days before a time shift versus the seven days after a time shift and (2) built a background distribution by making that same calculation for each day of the year. We then asked if time shift effects were outliers relative to the background. Several CC areas had non-zero time shift effects (Fig. 4A) but injury was the only area with a time shift effect in the top 15^th^ percentile (Fig. 4B). In sum, except for injury, our evidence for acute effects of time shifts is weak, especially when compared to the large weekly and seasonal effects on almost all types of pediatric emergencies.

**Fig 4.**
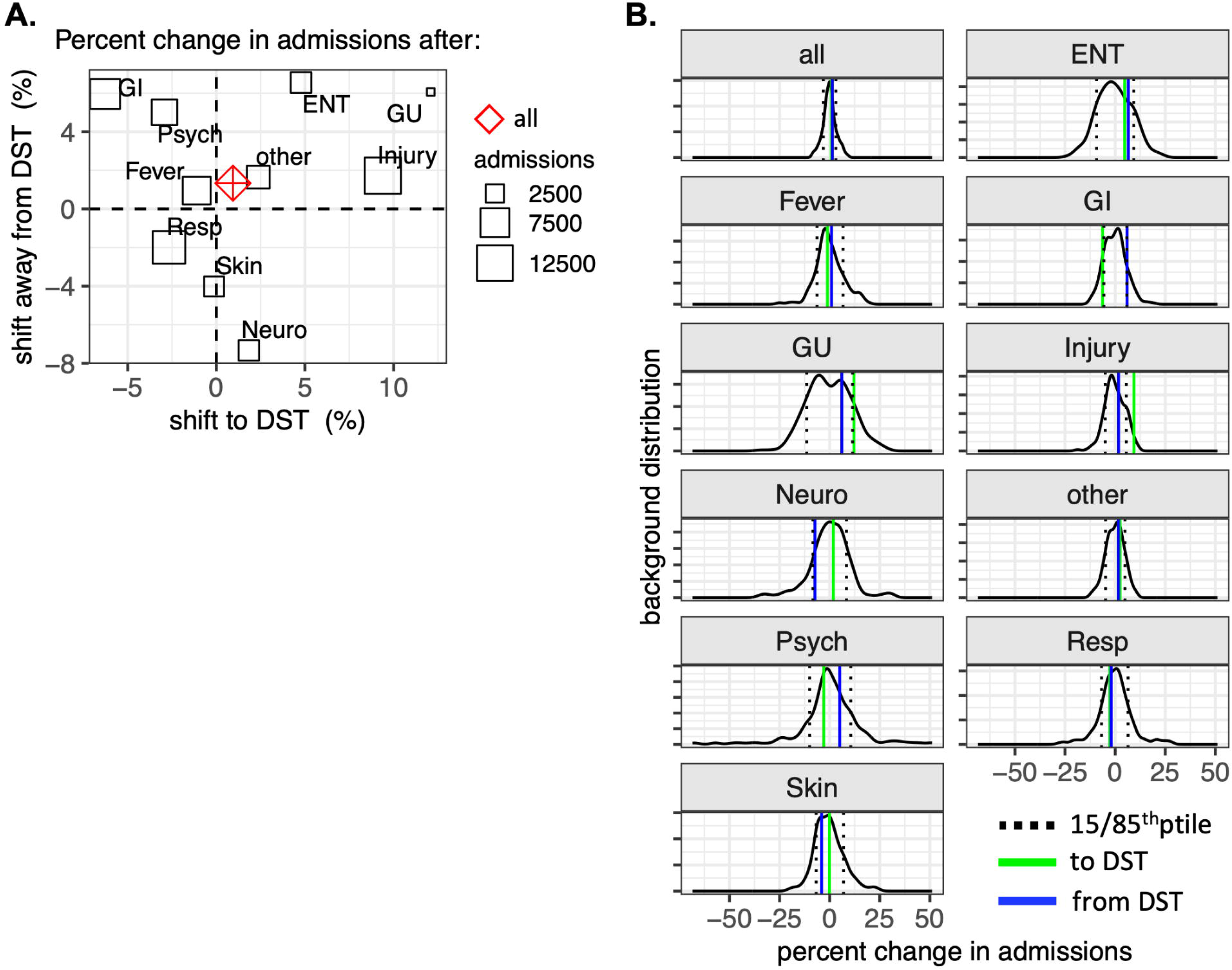
Acute impact of clock time shifts on injury admissions. **(A)** Percent difference in admissions the seven days before a time shift versus the seven days after a time shift. We evaluated shifts to (x-axis), and away from (y-axis) daylight savings time (DST). Positive values indicate increases in admissions in the seven days after a time shift, for overall admissions (red diamond) or CC areas. *ENT: ear, nose, throat; GI: gastrointestinal; GU: genitourinary; Neuro: neurological; Psych: psychiatric; Resp: respiratory.* **(B)** Background distributions built from the same calculation as in (A) but for each day of the year (n = 366).

### Annual rhythms of pediatric asthma- and psychiatric-related emergencies in Brazil

Annual asthma rhythms were observed in all four locations of Brazil studied (Fig. 5 and fig. S4). In each, the rhythms were unimodal, contrasting the bimodal patterns at our institution in Ohio, US (fig. S1). Porto Alegre, the location in the highest latitude–where seasonal variations in photoperiod and temperature are the highest–had the strongest seasonal effect with admissions 3.7 times higher than the trough. Recife, the location closest to the equator, had the lowest PTR at 1.4, followed by São Paulo and Brasilia. Asthma admissions peaked in winter in São Paulo and Porto Alegre, the southernmost locations. In Recife, admissions were steady year-round except for a decrease during the school summer break (December and January), suggesting the potential role of school exposures on asthma (26). In Brasilia, where there is a strong seasonality in precipitation, cases begin to rise in early summer (the wet season), peak in fall, and are the lowest during the dry winter months. Increased ED asthma admission during the wet season has been reported in other parts of the world, including Trinidad (27)and Mexico (28). Proposed causes include the increase in indoor allergen exposure and impact of humidity on pollen-related allergens(29).

**Fig 5.**
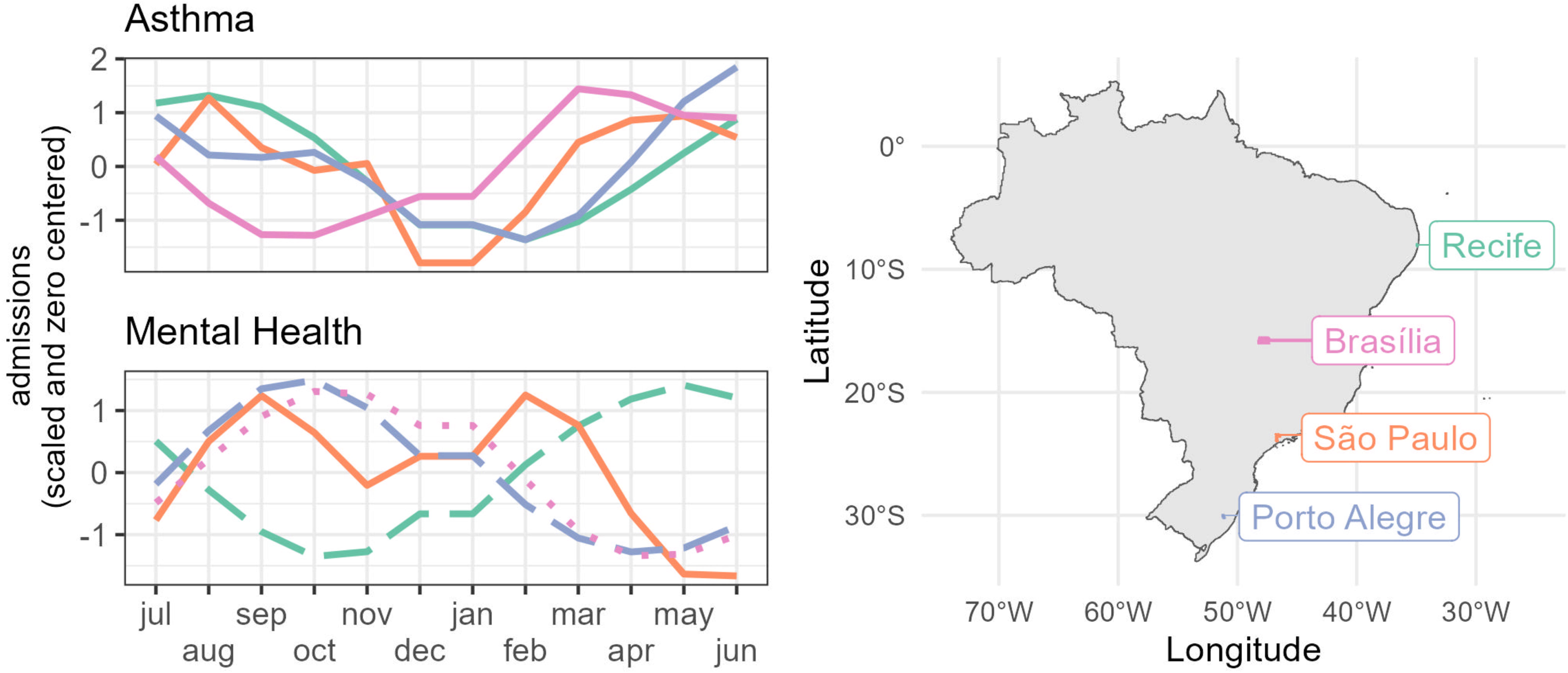
Annual patterns in asthma and mental health emergency/urgent admissions. *Left:* Scaled GAM smooth curves (see “Annual patterns of CCs” in Methods) of annual rhythms in asthma (top) and mental health (bottom) admissions. Each curve corresponds to one location: Recife (green), Brasilia (pink), São Paulo (orange) and Porto Alegre (blue). Line types indicate the model’s p-values: full (p<0.01), dashed (p<0.05), dotted (p >0.05). *Right:* Map of Brazil indicating the location of each chosen municipality.

We next evaluated psychiatric-related ED admissions (see Methods for cohort definition). Three of the locations had discernible annual rhythms: São Paulo, Recife and Porto Alegre (Fig. 5 and fig. S5). In São Paulo, a bimodal pattern was observed, with peaks after both winter and summer school breaks, similar to the pattern for the CC *Psychiatric Evaluation* at our institution in Ohio, US. Conversely, Porto Alegre and Recife were unimodal and peaked at opposite times of year. In Porto Alegre most psychiatric-related admissions occurred at the end of winter and beginning of spring. In Recife, admissions peaked during fall, before the school winter break. Interestingly, Recife had the highest effect size (PTR = 1.4), while both São Paulo and Porto Alegre had a PTR of 1.2. All effects sizes for mental health seasonality in Brazil were smaller than the ones found in Cincinnati, where most psychiatric conditions had a PTR above 2 (Fig. 1). The finding that the lowest-latitude location, with minimal seasonal changes in light and temperature, had the highest seasonal effect suggests a role for social factors in psychiatric-related ED admissions.

## Discussion

Approximately 70% of pediatric hospital admissions in the United States originate from the emergency department (33). The ability to predict patient ED visits is therefore crucial for alleviating overcrowding and optimizing care strategies. Various statistical models, as reviewed in (34), have been developed to forecast ED volume. These models, utilizing linear regression with calendar variables or time series techniques, can account for 31–75% of patient-volume variability. While the day of the week is consistently the most influential predictor, it only explains a portion of the variability. Importantly, these models forecast overall ED volume but do not account for specific medical conditions. We observed annual rhythms for a broad spectrum of chief complaints (CCs) in the ED, spanning *Head Injury, Migraine*, *Suicide Ideation*, *Skin Infection*, and many others. CCs peaked at different times of year, suggesting that accounting for type of conditions may help to best align resources with demand.

We compared rhythm effect sizes for all CCs, defined as the difference in the number of ED admissions between peak and trough times of the year. Notably, the CC *Psychiatric Evaluation* ranked among the largest effects, with a rhythm effect size on par with the CC *Fever* associated with wintertime infections. These findings are applicable beyond our institution and provide insights into key risk factors. A recent (2023) analysis by the Centers for Disease Control and Prevention (CDC) of ED visits from 1,919 facilities in 46 US states focusing on mental and behavioral health conditions among children aged 5–17 years old from 2018 to 2023, found a twofold increase in ED visits during the fall school semester, persisting throughout the spring semester (35). This aligns with our observations, and offers insights into how the ED and healthcare system might be organized around seasonality.

However, understanding patterns in mental health crises can be challenging, as individuals with a shared condition, or CC, also have many non-overlapping comorbidities and demographics. In fact, the annual curve for any defined group can be misleading if hidden patient types and rhythms exist. To study this, we used latent class analysis to cluster patients in each CC based on their discharge codes, and then compared the annual profiles of CC clusters. For example, patients with the CC *Psychiatric Evaluation*, the most prevalent psychiatric-related CC at our institution, had an overall rhythm with both spring and fall peaks. However, class modeling identified fall-peak, spring-peak, and arrhythmic classes, suggesting psychiatric phenotypes with distinct seasonal influences. All classes were enriched for the ICD-10 code for suicide ideation, but otherwise differed in ICD-10 composition, including different proportions of attention deficit hyperactivity disorder, intermittent explosive disorder, and other psychiatric and non-psychiatric diagnoses. Future studies using these informatic approaches can be extended to larger datasets and additional variables.

Geographic and socio-cultural variables are important to consider. Our study, along with the CDC report (35) and other US-based studies (8, 36–44), showed a higher frequency of pediatric mental health-related ED visits during school months. However, apart from a study in Japan that supports this school association (45), it remains uncertain whether this phenomenon is worldwide. Our analysis of ED data from Brazil suggests that the link between school and mental health crises in the US does not generalize globally. In fact, in several regions of Brazil, the relationship was the opposite, with school appearing to be a protective factor rather than a risk factor. The reasons for this contrast warrant further study, highlighting the complexity of seasonality in health.

Endogenous biological rhythms in physiology and behavior are important to consider. These rhythms are observed in various aspects of animal biology, such as morphology, migration, reproduction, and sleep-activity patterns (46). While the seasonality of human biology is less apparent, population and longitudinal studies demonstrate annual variations in immune (21), endocrine (20, 47, 48), cognitive (49), and other functions (50). Current understanding, primarily derived from animal models, suggests that organisms use circadian clock mechanisms to track changing day length, providing a time reference for the coming season (50–54). Although the role of endogenous seasonal clocks in human health remains uncertain, it is plausible that these clocks play a part in the annual patterns of illness and disease. Just as circadian biology explains increased morning risks of heart attack and stroke (55, 56), paralleling daily rhythms in physical activity, blood pressure, and coagulability (57, 58), the same concept could apply to seasonally triggered health conditions.

Alternatively, a condition may be triggered by disruptions in the seasonal clock. While this idea is speculative, disrupted circadian clocks, caused by sleep disturbances and shift work, have been linked to many health issues (57). Also, the duration and quality of human sleep may vary seasonally (59), and, in children, may be influenced by school (60). Sleep is associated with various medical conditions and health outcomes (61), potentially exacerbating mental health issues, migraines, injuries, and other conditions whose phases align with school schedules.

These areas warrant further investigation.

Our study has several important limitations. First, most of our findings are based on data from a single large tertiary care children’s hospital located in an urban area of the United States. This location, in a temperate zone, undergoes the full spectrum of annual changes in day length, temperature, precipitation, and vegetation. Consequently, the generalizability of our findings to other pediatric hospitals worldwide is not straightforward. While many of our findings, such as annual rhythms in mental health (45, 62), migraine (63), and injury (64), have been previously observed in various other settings, our analysis of ED visits in Brazil suggests potentially significant global variations. To address this, a unified framework is needed for modeling annual variations on a global scale.

A second limitation is our reliance on structured data within the EMR, including chief complaints and ICD-10 diagnosis codes. We did not cross-validate this information with other sources, such as medication usage or patient charts. While we assumed that the extensive patient dataset would mitigate random misdiagnoses, we acknowledge the potential for systematic biases in patient classifications that could influence our interpretation and subsequent generalizability.

A potential third limitation pertains to our treatment of ED visits as independent events, even though some individuals may have had multiple ED visits. We made this choice because CCs could vary between visits for the same individual, and we believed that repeated measures were unlikely to influence the overall annual patterns. However, this approach deviates from statistical convention, which assumes independence among all observations in population-level analyses. Future analyses could explore patient-level patterns, potentially revealing whether certain individuals are more prone to utilizing the ED at specific times of the year.

A fourth limitation concerns our selection of the number of patient classes to model for each CC. Our objective was to achieve maximum class homogeneity in ICD-10 profiles while still maintaining a sufficient number of admissions for annual assessment. However, for very few CCs, we were able to achieve complete homogeneity, resulting in patient groups that are similar but not identical. For many of these CCs, clustering more deeply may help to interpret patient and rhythm subtypes. Additionally, rule-based algorithms for grouping based on predefined clinical criteria may improve class formation.

## Materials and Methods

### Study cohort

Approval for this study was obtained from the Cincinnati Children’s Hospital Medical Center institutional review board. Consent was not required for this retrospective analysis of electronic medical records. All analyses were done in R language. Anonymized data and code to reproduce analyses are available at https://zenodo.org/records/14502802.

Of 1,653,828 total admissions to our institution’s emergency department and five affiliated urgent care centers from January 2010 to December 2021, we excluded admissions (i) without date-time information or (ii) with a subsequent admission within seven days for the same patient. This left 1,545,630 admissions (93% of total) of 501,687 patients for analysis.

### Annual patterns of CCs

To detect annual patterns of CCs, we used a generalized additive model (GAMs) framework as implemented in the mgcv package in R (65). Unlike other linear models, GAMs do not assume linearity. Instead, the effects of continuous predictors (eg. month) and categorical predictors (eg. biological sex) are described by spline functions (*s*) parametric functions (*f*), respectively. CCs with at least 500 admissions were binned into 12 monthly counts and modeled as a smooth annual curve, using the formula: Admissions ∼ *s*(month, basis = cc), where ‘cc’ = circular constraint. Global settings were ‘Identity’ = poisson; ‘Method’ = REML.

### Rhythm clusters

We used k-means clustering to partition the 244 CCs with annual variation into six groups based on their GAM-estimated smooth curves. The choice of six clusters was based on visual analysis of an “elbow” plot of the within-cluster sum of squares for k = 1 through k = 12 clusters. We identified six clusters as the point in the plot at which the rate of decrease in within-cluster sum of squares diminishes.

### Rhythm effect sizes

We assigned a peak and trough to each of the 244 rhythmic CCs as the average number of admissions in the top (peak) and bottom (trough) two months. These values were used to compute a peak-to-trough ratio and peak-minus-trough value for each CC.

### Covariate interactions

To test for potential interactions between (1) month and year, and (2) month and sex on admissions, we evaluated how well each CC was fit by four models, using the formulas: 1a. Admissions ∼ *s*(month) + *s*(year) [no interaction]

1b. Admissions ∼ *s*(month + year) [interaction]

2a. Admissions ∼ *s*(month) + *f*(sex) [no interaction]

2b. Admissions ∼ *s*(month, by = sex) + *f*(sex) [interaction]

Global settings were ‘Identity’ = poisson; ‘Method’ = REML. We used percent change in Akaike Information Criterion (pΔAIC) to assess improvement in fit when CC admissions were modeled as an interaction between month and year (or sex) versus no interaction. Positive pΔAIC values suggest the presence of an interaction between month and year (or sex), with larger values indicating stronger interactions. Top Interactions were inspected visually (fig. S2B&C).

### CC patient complexity

We randomly selected 5% of total admissions for each CC. Using the Jaccard index, we calculated the similarity in ICD-10 profiles between all possible pairs of admissions. The Jaccard index represents the intersection size divided by the union size of two binary vectors. The intersection size indicates the number of positions where both vectors have a value of 1 (indicating the presence of the corresponding ICD-10 code). The union size indicates the number of positions where at least one vector has a value of 1. Finally, we computed the mean of all Jaccard indices to obtain a single measure of similarity for each CC.

### Latent class analysis

We applied latent class analysis (LCA) to each CC with at least 1,000 admissions using the R package poLCA (67). For each of these 194 CCs, input to LCA was a data matrix with one row per admission and one column per ICD-10 indicating the presence or absence of that ICD-10. ICD-10s were excluded from analysis if they occurred in less than 1% of the CC population. In choosing the number of classes to fit to each CC, our goal was maximal class homogeneity while retaining enough admissions to evaluate seasonality. We initially fit four latent classes to each CC. If any classes had fewer than 250 admissions, LCA was rerun with N-1 classes, and so on until all classes had at least 250 admissions. Global settings in poLCA were = 10,000. 33 of the 194 CCs could not be split into multiple classes and so were excluded from subsequent between-class comparisons of annual variation.

### Spearman rank correlation

Spearman rank correlation, also known as Spearman’s rho, measures the strength and direction of the monotonic relationship between two variables, and can be useful for comparing the shapes of two different time-series profiles (68–70). In our study, each CC class was represented by a time-series with 12 values corresponding to the total admissions counts for each month of the year. We computed Spearman’s rho for each unique pair of classes within a CC, resulting in six rho values if there were four classes. A rho value of 1 indicates a perfect positive correlation, meaning that the two classes rise and fall at the same time. A rho value of -1 indicates a perfect negative correlation. From the pair values, we computed a mean rho value for each CC to provide a single measure of inter-class similarity in annual patterns of admission.

### Rolling change

We computed 3-day rolling rates of change in admissions over the full study period from Jan 01, 2010 to Dec 31, 2021. For each day of the year, the rate of change was defined as the difference between the last and first values in the 3-day rolling window divided by the first value. The rate of change plotted corresponds to the last value in each window.

### Data from Brazil

Data was extracted from the DATASUS database, which gathers data from all information systems of the public Brazilian Unified Healthcare System (SUS). We used the R package microdatasus(66) to extract microdata from the database. We focused on urgent and emergency hospital admissions of people between 0 and 18 years old. Data between 2012 and 2021 was pulled for four municipalities: Recife, Brasilia, São Paulo, and Porto Alegre. All municipalities chosen are capitals of their respective states and have a population above 1 million people. We considered asthma admissions where the main diagnosis ICD-10 code was J45. For mental health, we considered all admissions that had a main diagnosis code from ICD-10’s chapter V. Effect-sizes were calculated as described above.

## Supporting information

Effect sizes for 244 CCs with annual rhythm

Class ICD-10 makeup for all CCs

Admit class assignments for all CCs

Annual profiles for all CC classes

## Data Availability

Code for pulling Brazilian data and doing the analysis is available online at 10.5281/zenodo.14502801.
A subset of deidentified data will be available upon request to authors. The full dataset cannot be shared due to HIPAA restrictions.

https://zenodo.org/records/14502802

## Ethics

The Institutional Review Board of Cincinnati Children’s Hospital Medical Center waived ethical approval for this work (protocol 2020-0799).

## Acknowledgments

This paper is dedicated to Hector Wong.

