## Supplemental Figures for "Widespread annual rhythms in pediatric emergencies"

1. Division of Biomedical Informatics, and.
2. Division of Neurology, and.
3. Division of Pulmonary Medicine, and.
4. Division of Human Genetics, and.
5. Division of Critical Care Medicine, Cincinnati Children's Medical Center, OH, USA
6. Department of Otolaryngology-Head and Neck Surgery, University of Cincinnati School of Medicine, OH, USA.
7. Department of Pediatrics, University of Cincinnati School of Medicine, OH, USA

* Marc D Ruben

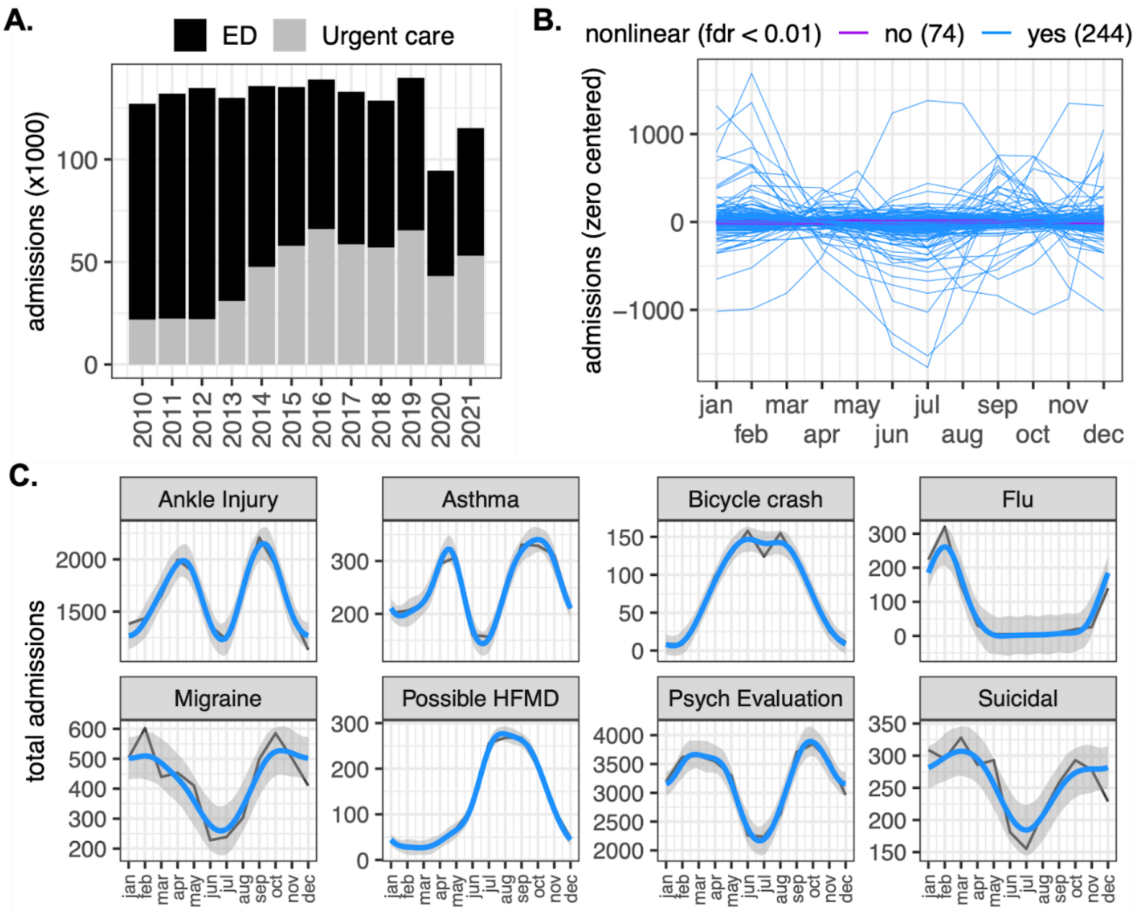


**fig. S1. Annual emergency department and urgent care admissions.** (**A**) Yearly admissions. (**B**) Monthly patterns of admission for each CC with at least 500 total admissions. This included 318 CCs from 1,103,199 admissions of 412,729 patients. For each CC, admissions were binned into monthly counts and modeled as a smooth annual curve by spline functions (see "*Annual patterns of CCs*" section in Methods). 244 of 318 (~80%) CCs were described by nonlinear patterns (FDR < 0.01). (**C**) Examples of rhythmic CCs. Black line: raw monthly counts. Blue line: GAM smooth annual curves, with confidence interval. *HFMD: hand foot mouth disease*.

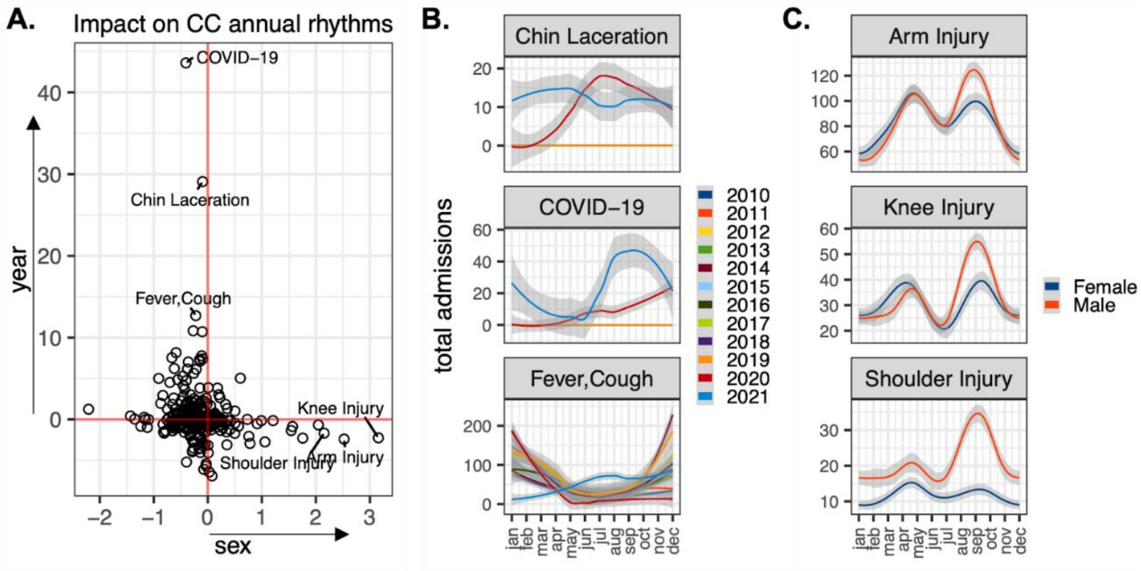


**fig. S2. Analysis of annual rhythms in relation to year and biological sex.** (**A**) Performance changes when modeling CC admissions as an interaction between month and sex (x-axis) or year (y-axis) compared to a model without interaction (refer to the "Covariate interactions" in Methods). Axes indicate the percent difference in Akaike Information Criterion (p$\Delta$AIC) where positive values indicate an interaction between month and sex or year. Labels indicate the top three interactions. (**B**) The impact of month on CC *Covid-19* admissions depends on the year because this CC did not exist before 2020. Likewise, monthly patterns in *Chin Lacerations* depend on the year because, for reasons unknown, this CC also did not exist prior to 2020. Monthly patterns in *Coughing–Fever* depend on the year, due in large part to deviation in 2021 from usual patterns. (**C**) Sex has a small but discernible influence on the annual rhythms of several injury-related CCs. For most CCs, the relationships between month and admissions were minimally affected by sex or year.

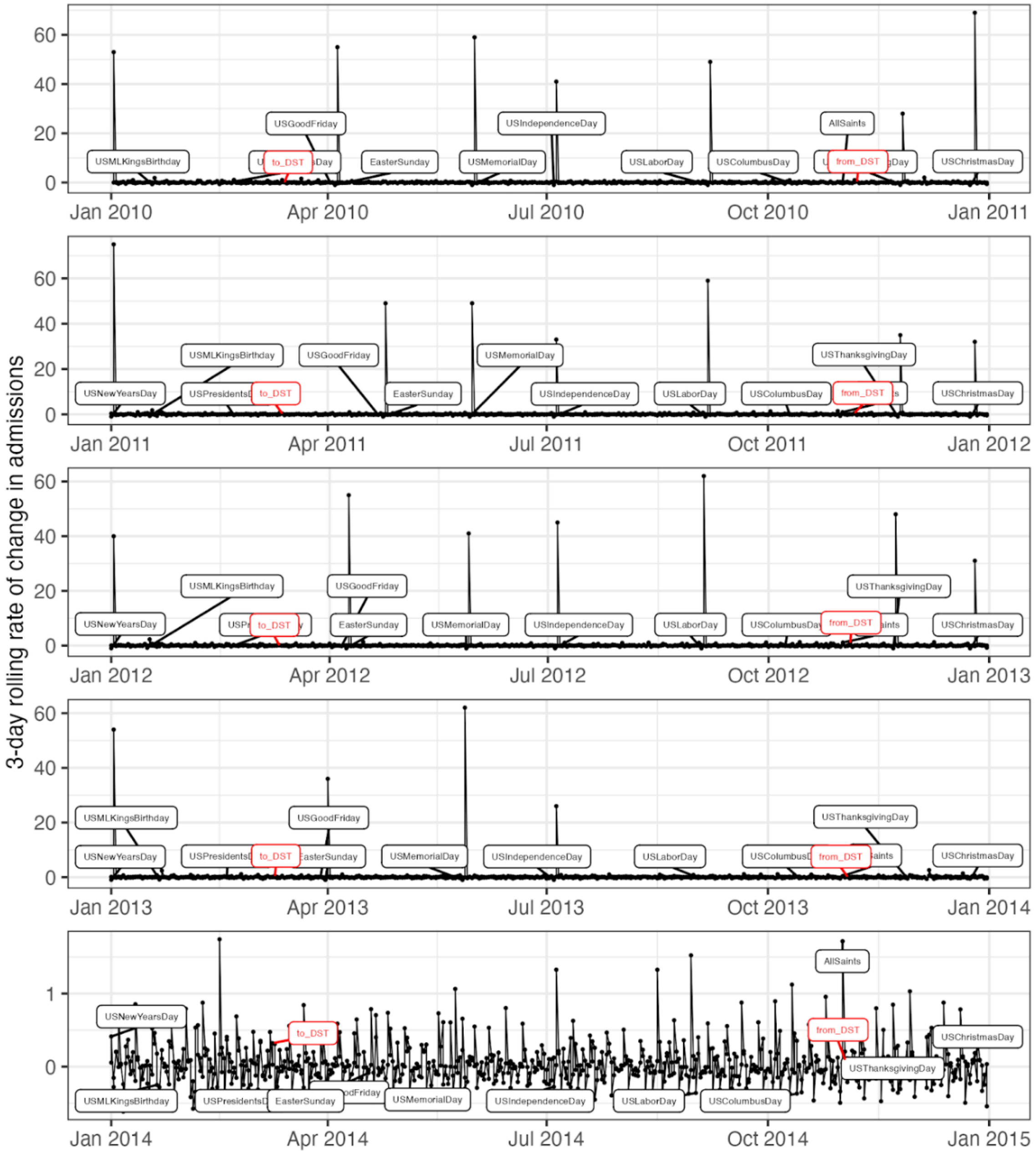


**fig. S3. Rolling rate of change in admissions from Jan 01, 2010 to Dec 31, 2014.** 3-day rolling rates of change in admissions from Jan 01, 2019 to Dec 31, 2020 (see “Rolling change” in Methods). Labels indicate major US holidays (black) and clock time shifts to and from daylight savings time (red). From 2010 through 2013, admissions appear to spike on the day after major US holidays. This is an artifact of our analysis because prior to 2014 UC clinics were closed on holidays, yielding spikes in the rolling rate of change for the day after. From 2014, UC clinics began remaining open on holidays and thus there were no obvious holiday effects. 2015–2021 are not shown for space reasons but the patterns in these years are consistent with data from 2014.

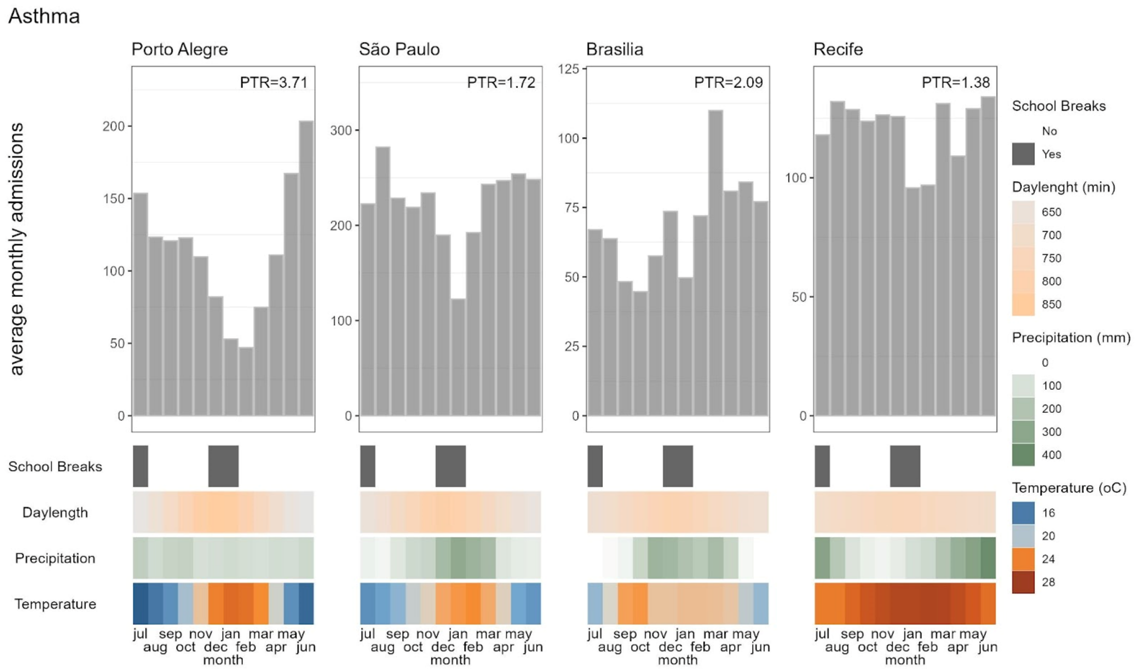


**fig S4.Average monthly asthma emergency admissions and monthly mean values in environmental variables at each of the four Brazilian location**s. Top: average monthly emergency asthma (ICD-10 J45) admissions in each municipality, calculated with data pulled from the DATASUS database from Jan. 2012 to Dec 2021. Bottom: Graphic representation of annual changes in social and environmental variables in each municipality. School breaks are represented as yes/no in the months they are most likely to occur; small variations in school districts and year-to year can be present. Daylength is the number of minutes between sunrise and sunset at the 15th of each month (data from https://aa.usno.navy.mil/). Precipitation and temperature are monthly means calculated with data from 1990 until 2020 (data from https://portal.inmet.gov.br/).

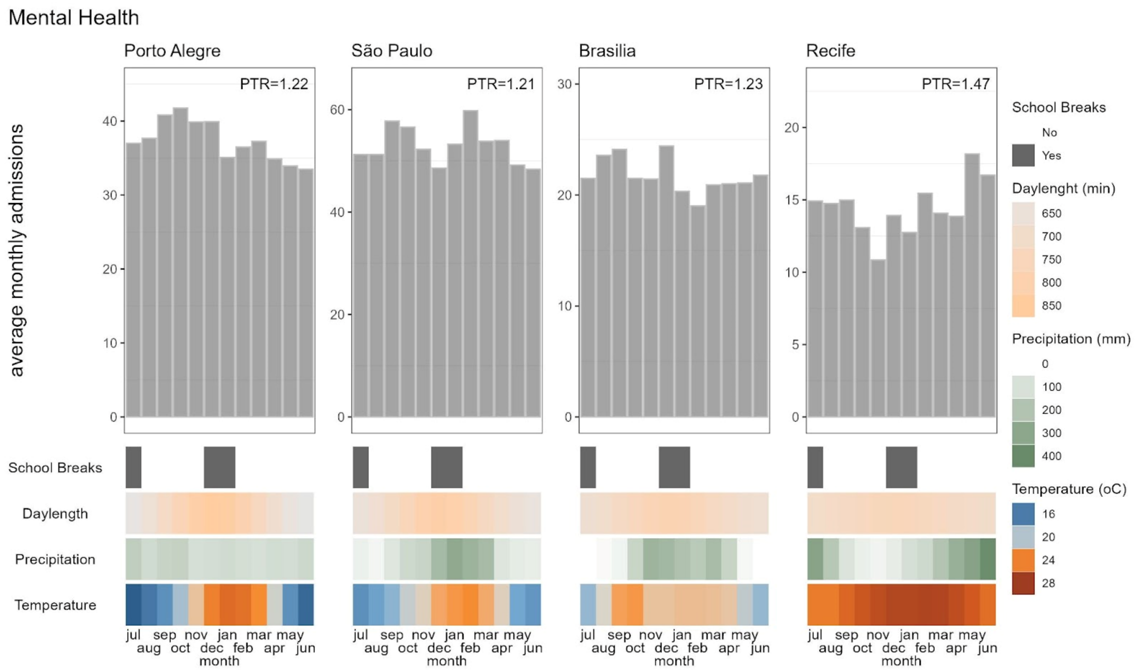


**fig S5.Average monthly mental health emergency admissions and monthly mean values in environmental variables at each of the four Brazilian location**s. Top: average monthly emergency mental health (ICD-10 Chapter V) admissions in each municipality, calculated with data pulled from the DATASUS database from Jan. 2012 to Dec 2021. Bottom: Graphic representation of annual changes in social and environmental variables in each municipality. School breaks are represented as yes/no in the months they are most likely to occur; small variations in school districts and year-to year can be present. Daylength is the number of minutes between sunrise and sunset at the 15th of each month (data from https://aa.usno.navy.mil/). Precipitation and temperature are monthly means calculated with data from 1990 until 2020 (data from https://portal.inmet.gov.br/).

**Supplementary Files**

**file S1. Effect sizes for 244 CCs with annual rhythms.**

**file S2. Class ICD-10 makeup for all CCs.**

**file S3. Admit class assignments for all CCs.**

**file S4. Annual profiles for all CC classes.**
